# Donor side effects experienced under minimal controlled ovarian stimulation (COS) with in vitro maturation (IVM) versus conventional COS for *in vitro* fertilization (IVF) treatment

**DOI:** 10.1101/2024.03.28.24304995

**Authors:** Maria Marchante, Ferran Barrachina, Sabrina Piechota, Marta Fernandez-González, Alexa Giovannini, Trozalla Smith, Simone Kats, Bruna Paulsen, Eva González, Virginia Calvente, Ana Silvan, Baruch Abittan, Joshua Klein, Peter Klatsky, Daniel Ordonez, Christian C. Kramme

## Abstract

**Objective:** To evaluate how minimal controlled ovarian stimulation (COS) for in vitro maturation (IVM) affects subjects’ oocyte retrieval experiences compared to conventional COS, considering side effects

**Design:** Retrospective Survey Study

**Setting:** Clinical in vitro fertilization (IVF) treatment centers in Spain and the United States.

**Subjects:** Data were collected from subjects undergoing minimal COS (n=110; 600-800 IU FSH) for IVM and conventional COS for egg donation (n=48; 2000-3000 IU FSH) from April 2022 to November 2023. In the same period, a pairwise comparison of subjects (n=13) undergoing both minimal COS for IVM and conventional COS for oocyte cryopreservation was conducted.

**Intervention/Exposure:** Minimal and conventional controlled ovarian stimulation.

**Main Outcome Measures:** The most common side effects suffered during ovarian stimulation and after OPU, satisfaction level, and the likelihood of recommending or repeating minimal or conventional COS. Statistical analysis included Mann Whitney, Wilcoxon, Chi-square, and McNemar tests, with a significance level set at p<0.05.

**Results:** During minimal COS, most subjects did not experience breast swelling (86%), pelvic or abdominal pain (76%), nausea or vomiting (96%), and bleeding (96%). After oocyte pick-up, the majority (75%) reported no pelvic or abdominal pain. The most common side effect was abdominal swelling (52%). Compared to conventional COS cycles, minimal COS subjects reported significantly less post-retrieval pain, with 33% experiencing no pain (vs. 6%; p=0.0011) and with a reduced severe level of pain (5% vs.19%; p=0.0097), leading to fewer subjects requiring pain medication (25% vs. 54%; p=0.0003). Additionally, 85% of women were very satisfied with minimal stimulation and would recommend or repeat the treatment. In the comparison in which each donor underwent both minimal and conventional COS treatments, women indicated more side effects with the conventional stimulation, presenting a significantly overall higher level of pain (p=0.0078).

**Conclusion:** Reducing the hormonal dose for ovarian stimulation has a beneficial effect on subjects, suggesting the combination of minimal COS with IVM techniques is a well-tolerated alternative for women who cannot or do not wish to undergo conventional controlled ovarian hyperstimulation.

## Introduction

Assisted reproduction treatments typically involve the administration of gonadotropins to stimulate the growth of multiple follicles in the ovaries, facilitating the retrieval of mature eggs for *in vitro* fertilization (IVF) (1). Controlled ovarian stimulation (COS) represents the conventional hormonal protocol, and involves the administration of high doses of gonadotropins throughout 9 to 14 days. Moreover, in addition to the discomfort of frequent and painful hormonal injections over this timespan, the COS hormonal regimen is associated with health side effects, significantly inflates IVF costs, and extends treatment duration, often creating barriers for patient access and utilization.

A major risk of conventional COS is the development of Ovarian Hyperstimulation Syndrome (OHSS). The risk for which is primarily correlated with ovarian reserve, number of oocytes retrieved, and the triggering agent utilized (2,3). Mild OHSS is estimated to occur in up to one in three women, and moderate to severe OHSS in 2-6% of cycles, with significantly higher risks for women with polycystic ovarian syndrome (PCOS) and predicted hyper responders (4–9). Recent improvements in monitoring as well as use of antagonists during stimulation and agonists for triggering have significantly lowered the risk of severe OHSS. However, mild to moderate cases still persist (10). Beyond OHSS, hormonal medications in conventional COS have a high prevalence of symptoms like mood swings, hot flashes, headaches, and bloating. The treatment may induce emotional and physical stress with frequent injections, monitoring appointments, and uncertainty in IVF success (11–13). Moreover, the high doses of hormones used in infertility treatments result in expensive costs for women who wish to preserve their fertility or achieve their desired motherhood, which often requires the need for multiple cycles (14). The health and financial consequences of conventional COS contribute highly to limiting overall access to IVF treatment and continue to reinforce inequality in care worldwide, which is a major concern globally for health equity in fertility treatment.

An alternative treatment to the conventional COS is minimal or abbreviated COS protocols paired with *in vitro* maturation (IVM) of oocytes, which substantially reduce gonadotropin use from 9-14 days to 2-4 days. This reduction in the overall gonadotropin dosage minimizes painful injections, decreases monitoring appointments, cuts medication costs, and reduces side effects and OHSS risk compared to conventional COS (15). However, while reducing the gonadotropins dosage shortens the stimulation process, typically, all retrieved oocytes are immature and require an IVM step to be used for IVF purposes (3). Indeed, in recent years there has been a growing interest in IVM and the maturation of oocytes outside of the body, a process which has been demonstrated by several groups worldwide (16–22). Importantly, in 2021, the American Society for Reproductive Medicine (ASRM) declared IVM as a non-experimental technique (23). Therefore, minimal COS followed by IVM can streamline and enhance the safety of the IVF process. Despite this, a comprehensive analysis comparing side effects associated with minimal COS for IVM versus conventional COS is still lacking, particularly across centers from different nations with significant practice variation. Furthermore, limited studies have assessed side effects outcomes in women who underwent both minimal COS and conventional COS in the same center, which would be highly informative given the subjective nature of pain.

Our group recently reported the use of minimal COS in combination with a novel IVM treatment containing hiPSC-derived ovarian support cells (OSC-IVM), demonstrating an improvement in the oocyte maturation rates and embryo formation rates compared to a traditional IVM treatment (22). This research study was conducted at fertility centers located in both the United States and Spain. Here, we sought to analyze side effects experienced during and after stimulation in subjects that underwent minimal COS compared to a site-matched cohort undergoing conventional COS. Furthermore, in a limited sample set, we analyzed side effects in subjects who underwent both minimal COS and conventional COS in the same study center, ensuring a highly controlled pairwise comparison of outcomes. We further compared efficacy outcomes of minimal versus conventional COS using publicly available data from recent randomized control trials (RCTs) to better contextualize the outcomes of the treatment approaches.

## Material and Methods

### Type of study

This study is a retrospective survey study aimed to evaluate the side effects suffered during minimal COS treatments performed to obtain immature oocytes for IVM. In addition, these symptoms were compared with the side effects derived from conventional COS cycles for obtaining mature oocytes for egg freezing, intracytoplasmic sperm injection (ICSI), and IVF cycles.

### Study groups

Data was retrospectively collected from research subjects who underwent minimal COS or conventional COS between April 2022 and November 2023 at Ruber Juan Bravo University Hospital, Eugin Group (Madrid, Spain), Spring Fertility (New York, USA), and Extend Fertility (New York, USA) with informed consent and ethical approval from CNRHA 47/428973.9/22 (Spain) and Western IRB No. 20225832 (USA), respectively. This research adhered to the ethical principles outlined in the Declaration of Helsinki.

Two sets of comparisons were performed:

**Data Set 1** (*Minimal COS with IVM versus site-matched conventional COS subjects in Spain*): Subject symptom surveys were collected from study subjects who underwent minimal COS for an IVM research study at Ruber Clinic in Spain. This IVM study was carried out on 156 subjects. 110 survey responses were obtained from the total 156 participants (70.5% of the cases). Moreover, information on pain levels and medication used were also collected from women who attended the clinic during the same period for egg donation after undergoing a conventional COS protocol and voluntarily responded to the Ruber Juan Bravo University Hospital pain assessment survey (n=48). Donor demographic characteristics including donor’s age, body mass index (BMI), hormone levels, antral follicle count (AFC) and oocytes retrieved for each group are presented in Table 1.

**Table 1.** Donor demographic characteristics including donor’s age, body mass index, hormone levels, antral follicle count and oocytes retrieved for each group.

|  | Data Set 1<br>Minimal COS | Data Set 1<br>Conventional COS | Data Set 2<br>Minimal COS | Data Set 2<br>Conventional COS |
| --- | --- | --- | --- | --- |
| **Sample size (n)** | 110 | 48 | 30 | 30 |
| **Age at retrieval (years)** | 27.3 ± 4.1 | 26.3 ± 4.4 | 31.4 ± 2.6 | 31.4 ± 2.6 |
| **BMI (Kg/m<sup>2</sup>)** | 23.5 ± 3.1 | 22.0 ± 2.8 | 24.3 ± 4.7 | 24.3 ± 4.7 |
| **AMH (ng/mL)** | 4.1 ± 2.5 | 4.2 ± 2.2 | 5.8 ± 3.2 | 5.8 ± 3.2 |
| **AFC (n)** | 19 ± 6 | 29 ± 12 | 28 ± 12 | 28 ± 12 |

COS, controlled ovarian stimulation; BMI, body mass index; AMH, anti-Müllerian hormone; AFC, antral follicle count.

**Data Set 2** (*Minimal COS with IVM versus conventional COS subjects undergoing both treatments in the United States*): In this cohort, the subject symptom survey data set was a pairwise comparison of study participants who underwent both minimal COS for an IVM study and conventional COS for social oocyte cryopreservation within three months. The questionnaire was provided to 30 women, obtaining responses from 13 (43%). This data was collected from Spring Fertility and Extend Fertility in the United States. Participant demographic characteristics including subject’s age, BMI, hormone levels, AFC and oocytes retrieved are presented in Table 1.

### Minimal and conventional stimulation protocols at the reproductive clinics

**Data Set 1** (*Minimal COS with IVM versus site-matched conventional COS subjects in Spain*):

Briefly, 156 high ovarian reserve subjects underwent minimal COS that started on the second day of the menstrual cycle with single injections over 3-4 days of recombinant follicle stimulating hormone (rFSH) (600-800 IU total, Ovaleap) followed by no trigger or a trigger of hCG (5,000-10,000 IU, Ovitrelle) when one or more follicle reached 10mm in size, with no follicles above 12mm. In parallel, 48 women attended the clinic for oocyte donation, undergoing conventional COS based on 9-12 days of rFSH (1,800-2,400 IU Total) followed by a trigger injection (5,000-10,000 IU hCG) when at least one follicle was 18 mm (Figure 1).

**Figure 1.**
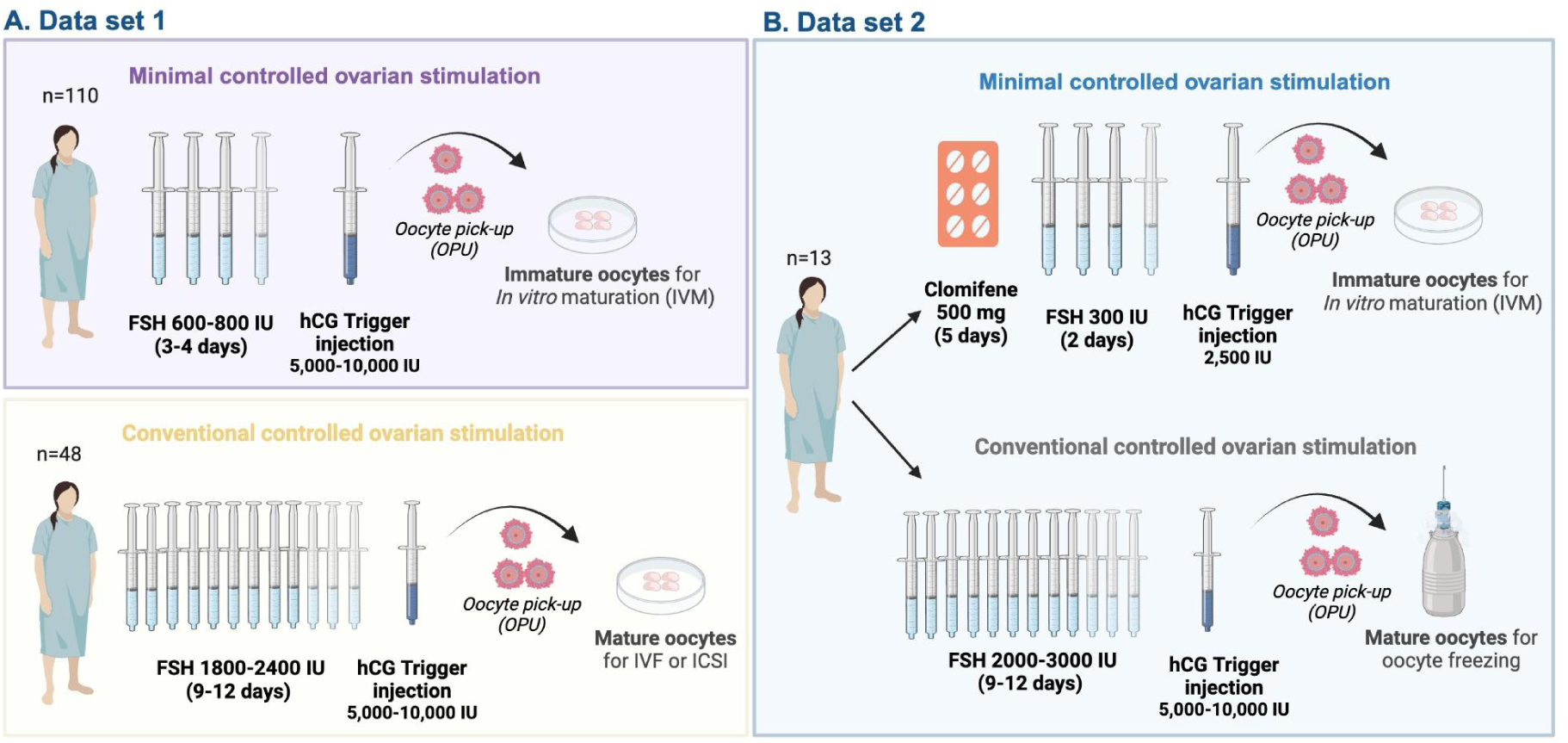
Schematic representation of the stimulation protocols. (A) Data Set 1. Minimal controlled ovarian stimulation (COS) for in vitro maturation (IVM) and conventional COS for in vitro fertilization (IVF) or Intracytoplasmic sperm injection (ICSI). (B) Data Set 2. Minimal COS for IVM and conventional COS for oocyte freezing. Follicle stimulating hormone (FSH); Human chorionic gonadotropin (hCG); International Unit (IU). Illustration created with BioRender.com.

**Data Set 2** (*Minimal COS with IVM versus conventional COS in subjects and social egg freezers undergoing both treatments in the United States*): In Data Set 2, 30 women initially underwent minimal COS and oocyte donation for an IVM research study, followed by conventional COS for social oocyte cryopreservation three months later, which was offered as a benefit for participating in the IVM research study. Starting on the second day of the menstrual cycle, the minimal COS comprised 5 days of oral clomiphene citrate (500 mg) and 2 days of injected rFSH (300 IU total) followed by no trigger or a trigger of hCG (2,500 IU) when one or more follicles reached 10mm in size, with no follicles above 12mm. Within 3 months of the minimal COS treatment, the same subjects underwent conventional COS based on 9-12 days of rFSH (2,000-3,000 IU Total) followed by a trigger injection (5,000-10,000 IU hCG) when at least one follicle was 18 mm (Figure 1).

### Follicle aspiration procedures for minimal COS and conventional COS

Oocyte retrievals were performed 34-36 hours after the trigger injection (2,500-10,000 IU hCG) or 48 hours after last rFSH injection for untriggered cycles using a transvaginal ultrasound with a needle guide on the probe to retrieve immature oocytes for IVM in the minimal COS treatments. Briefly, oocyte pick-ups (OPU) were done without follicular flushing using single lumen 17- or 19-gauge needles. Vacuum pump suction (70-140 mm Hg) was used to harvest follicular contents through the aspiration needle. In all cases, rapid rotation of the aspiration needle around its long axis when the follicle had collapsed provided a curettage effect to assist the release of COCs into the aspirate fluid. Although follicles were not flushed, the aspiration needle was removed from the subject and flushed frequently throughout the oocyte retrieval procedure to limit clotting and needle blockages. Aspirations of conventionally stimulated subjects were performed with 17-gauge needles, and 120 mm HG vacuum pump suction. Minimal curettage was utilized, and no follicular flushing was performed in conventional COS treatments.

After oocyte retrieval, follicular aspirates were examined in the laboratory using a dissecting microscope. Oocytes from the minimal stimulation cycles were used for further IVM experiments, while oocytes from the conventional were used for IVF or oocyte freezing.

### Collection of subject side effect data

A subject symptom and side effects survey was created with the goal of assessing various common side effects and overall experience of the stimulation protocol, retrieval procedure, and post-retrieval procedure.

The survey questionnaire was created based on the most common side effects after conventional ovarian stimulation (24) by fertility doctors (obstetrician/gynecologists and reproductive endocrinologists) following the construction guidelines for research questionnaires (25–26). Among the main side effects that subjects were surveyed about and self-reported were breast and abdominal swelling, nausea or vomiting, and pelvic/abdominal pain during hormonal injections. In addition, pelvic/abdominal pain and the medication needed immediately after OPU and 8-10 hours later were examined. The questions about pain levels and analgesia use were based on a standard questionnaire created and approved by the ethical and scientific committee of the Hospital Ruben Juan Bravo for pain assessment, aimed at capturing valuable healthcare insights, and to be used for teaching and research. The degree of pain was measured using a standard numeric rating scale from 0 (no pain) to 10 (the most pain).

As secondary outcomes, the timely resumption of their normal menstrual cycle and unexplained fever episodes or bleeding were also recorded. Finally, subjects were asked about their satisfaction level after oocyte donation under minimal COS treatment and if they would recommend or repeat the minimal COS donation with the level of personal satisfaction measured from 0 (not satisfied) to 10 (very satisfied).

For the subjects who underwent only minimal COS or conventional COS (Data Set 1), the questionnaires were conducted by telephone and using physical forms within one month after treatment. A web-based survey was employed for the participants who underwent both minimal COS plus IVM and conventional COS for oocyte cryopreservation (Data Set 2) after finishing both treatments. The same survey questions were used for minimal and conventional stimulation to ensure accurate comparison in Data Set 2.

The questionnaire included the possible answers for each question to facilitate the response and minimize human error. In the case of scale use, the score meanings were also written. When the surveys were answered by telephone or physical forms, the clinical staff reviewed them. For web-based surveys, subjects could fill them out just one time before sending them, avoiding multiple answers. In addition, to prevent unanswered responses, the web-based survey version was designed to require marking an option before proceeding to the next question. In both sets of experimental comparisons, subject responses were kept anonymous. The clinic coordinators managed the data, creating an anonymized code for each donor interviewed, ensuring data privacy and confidentiality at the different clinics. The questionnaire was presented to the clinical staff before conducting the survey to clarify any possible questions.

### Statistical analysis

Data analyses were conducted using GraphPad Prism software (GraphPad Software, San Diego, CA, USA). Categorical variables were described as absolute numbers and percentages. Continuous variables were described on a scale of 0 to 10. A power calculation was not performed as this was a retrospective study, and the sample size was derived from the available clinical database. For statistical analysis of pain distribution after OPU for Data Set 1, data was analyzed via two-tailed Mann Whitney U-Test. For pain distribution after OPU for matched samples in Data Set 2, data was analyzed via two-tailed, Wilcoxon matched-pairs sign rank test. For categorical variables in unpaired data (Data Set 1), a two-tailed Chi-square test was utilized. For categorical variables with paired data (Data Set 2), McNemar’s test was utilized. A *p*-value of less than 0.05 was considered statistically significant in all tests.

## Results

### Side effects experienced during minimal COS treatment

From study subjects who underwent minimal COS for an IVM research study at Ruber Clinic, we were able to obtain side effect responses from 110 of 156 subjects. This represents 70.5% of the IVM study population. First, we analyzed the side effects experienced during hormonal injections, observing that subjects under minimal COS treatment reported a low prevalence of side effects commonly experienced in conventional COS treatments. Specifically, most minimal COS subjects did not suffer breast swelling, pelvic or abdominal pain, nausea, or vomiting during the minimal stimulation. However, around half of the subjects did experience abdominal swelling, which was the most reported side effect during minimal stimulation (Table 2).

**Table 2.** Side effects experienced during minimal COS treatment.

| During Minimal Stimulations were the following experienced? (N=110) |  | Presence of symptoms |
| --- | --- | --- |
| During Stimulation | Breast swelling | 16 (14.55%) |
|  | Abdominal swelling | 57 (51.82%) |
|  | Pelvic or abdominal pain | 26 (23.64%) |
|  | Nausea or vomiting | 5 (4.55%) |
| Post OPU | Pelvic or abdominal pain | 27 (24.55%) |
|  | Pain medication needed* | 27 (24.55%) |
|  | Bleeding | 4 (3.64%) |
|  | Fever | 2 (1.82%) |
|  | Regular menstrual cycle after a month | 100 (90.91%)** |

\*Ibuprofen, paracetamol; \*\* 5 (4.55%) subjects had previously irregular menstrual cycles

After oocyte pick-up, the majority (75%) of subjects reported having no pelvic or abdominal pain, and pain medication like paracetamol or ibuprofen was not needed. Most subjects (96-98%) did not suffer bleeding or fever because of the oocyte retrieval. Moreover, the vast majority of subjects resumed their regular menstrual cycles the following month after the minimal stimulation treatment. Among the limited number of subjects who did not resume regular cycles (∼9%), half had a prior history of irregular menstrual cycles (Table 2).

### Levels of pain under minimal stimulation compared to conventional stimulation

Subjects who underwent minimal COS experienced less pain after OPU compared to subjects who underwent conventional COS treatment. Comparison of pain levels experienced under the minimal stimulation versus the conventional showed that 8 to 10 hours after OPU, a significantly higher percentage of subjects reported no pain (level=0) in the minimal stimulation group (32.73%) as opposed to the conventional COS group (6.25%) (p=0.0011, Chi-square, two-tailed) (Figure 2A). Low and moderate levels of pain (level 1-3 and level 4-7 respectively) were lower but not significantly in the minimal stimulation group (36.4% and 26.4%) compared to the conventional COS group (39.6% and 35.4%) (p= 0.8368 and 0.3362, Chi-square, two-tailed) (Figure 2A). The effect of stimulation dosage was especially striking in levels of severe pain, where the percentage of subjects in the conventional COS group increased significantly over threefold compared to the minimal stimulation group (18.8% versus 4.6%) (p=0.0097, Chi-square, two-tailed) (Figure 2A). Comparison of the overall magnitude of pain distribution between groups showed that subjects who underwent minimal stimulation experienced statistically significantly less pain after OPU compared to the conventional COS treatment group (p<0.0001, Mann-Whitney U-Test, two-tailed) (Figure 2B). When the pain levels warranted it, subjects took medication 8 to 10 hours after oocyte pick up. As expected, significantly less subjects under minimal stimulation compared to conventional COS required medication for pain management (24.55% versus 54.17%, p = 0.0003, Chi-square Test, two-tailed) (Figure 2C).

**Figure 2.**
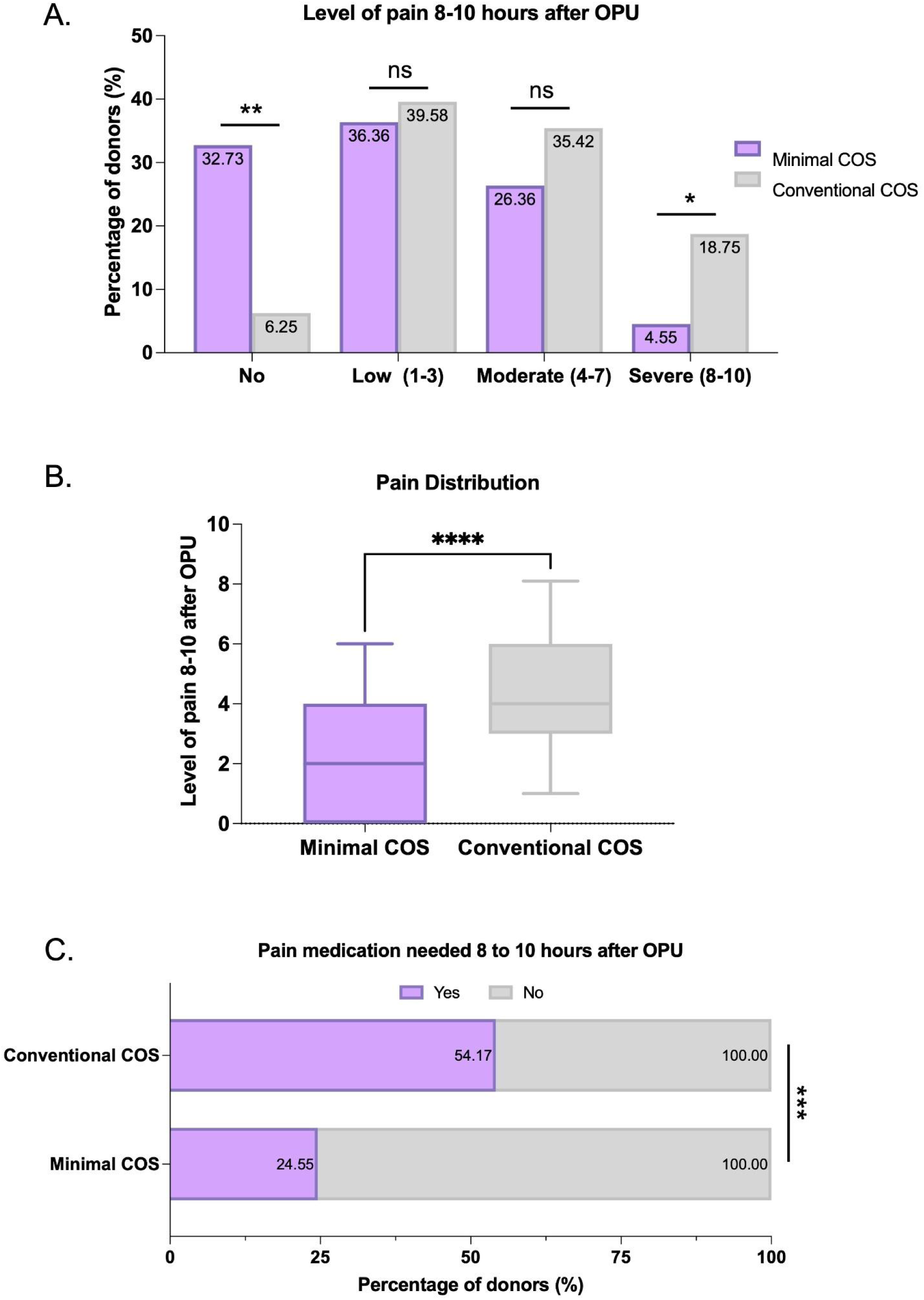
Pain experienced during minimal versus conventional ovarian stimulation. (A) Percentage of subjects experiencing each level of pain 8 to 10 hours after oocyte pick up (OPU) in the minimal controlled ovarian stimulation (COS) and conventional COS groups. Statistical analysis for each pain range (none, low, moderate, severe) was performed via Chi-square test with Yates correction, two-tailed, *, p=0.0097, **, p=0.0011 (B) Distribution of pain values in both conditions on a scale of 0-10, data is plotted as a box and whisker plot, with a median and a 90% confidence interval. Statistical analysis was performed via Mann-Whitney U-Test, two-tailed. ****, p<0.0001. (C) Percentage of subjects who needed pain medication 8 to 10 hours after OPU. Statistical analysis performed via Chi-square test without Yates correction, two-tailed, ***, p= 0.0003.

### Satisfaction level after oocyte donation with minimal COS

After the minimally stimulated donation process for IVM, subjects were asked about their level of satisfaction. 84.55% of women indicated they were very satisfied with the treatment, and 5.45% were satisfied, and no donor reported being unsatisfied (Figure 3A). Overall, 86.40% of subjects indicated that they would repeat or recommend a minimal stimulation treatment (Figure 3B).

**Figure 3.**
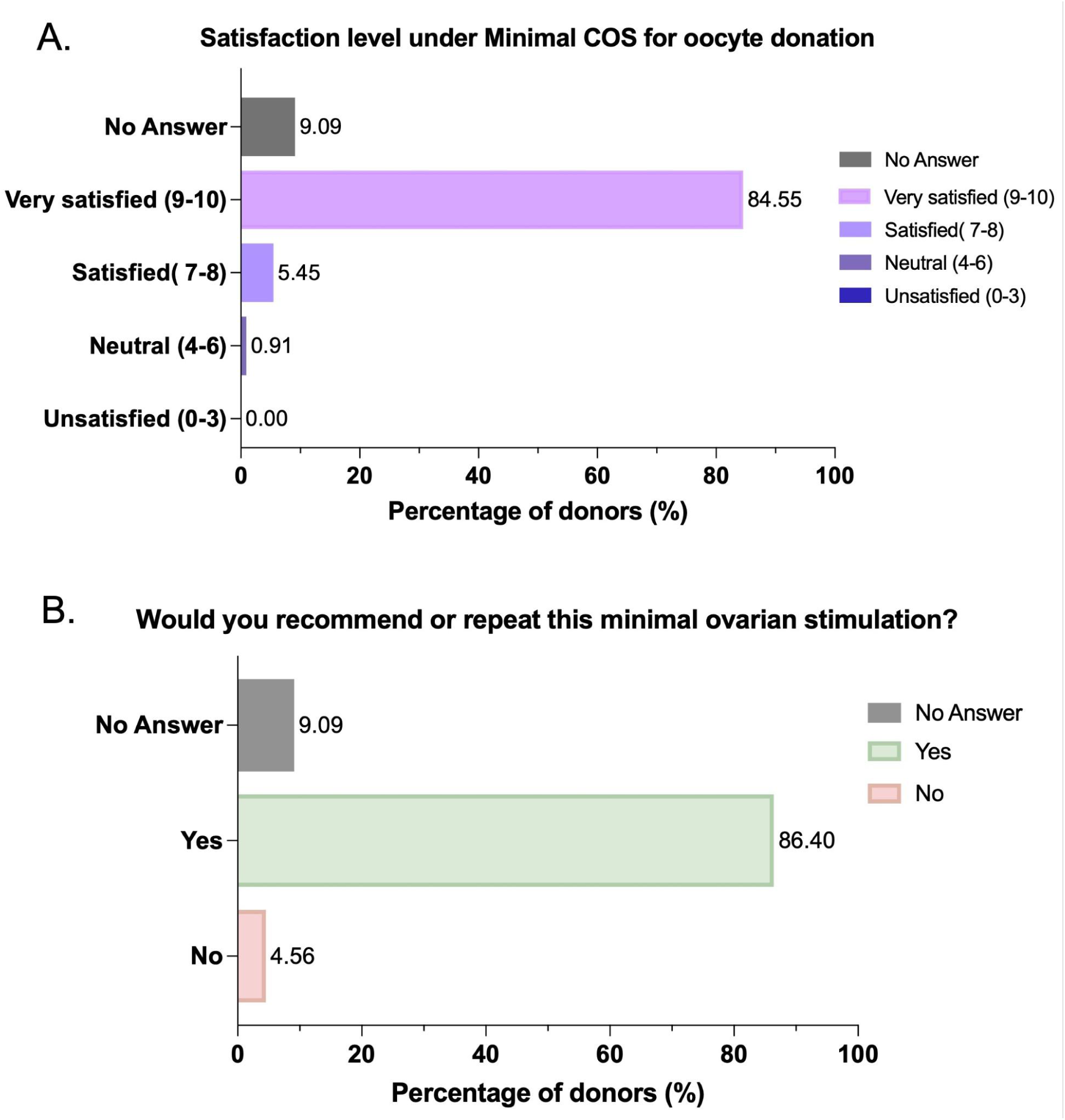
Satisfaction after oocyte donation with minimal stimulation. (A) Satisfaction level classified by stimulation group from very satisfied to unsatisfied, measured on a scale from 0 to 10. (B) Percentage of subjects who would/would not recommend or repeat the minimal stimulation treatment.

### Pairwise comparison of women who underwent both minimal and conventional hormonal stimulation

Thirteen responses were submitted from the thirty participants who underwent both stimulation types within three months at the same center in the United States. That represents 43% of the total subjects, considering the number of participants that fully completed the survey divided by the total number of participants presented with the questionnaire. Overall, there was a trend towards participants indicating more side effects were experienced after conventional COS compared to minimal COS, with more instances of pain, swelling, nausea, and vomiting (Table 3). Post-retrieval, women experienced more pain, usage of pain medication, and bleeding after conventional COS compared to minimal COS, while similar levels of nausea, vomiting, or fever were noted between the regimens (Table 3).

**Table 3.** Symptoms experienced by patients from minimal and conventional stimulations.

| During Minimal & Conventional Stimulations were the following experienced? (N=13) |  | Minimal Stimulation | Conventional Stimulation | p-val |
| --- | --- | --- | --- | --- |
|  |  | Yes |  |  |
| During Stimulation | Breast swelling | 4 ( 30.77%) | 6 (46.15%) | 0.48 |
|  | Abdominal swelling | 8 (61.54%) | 11 (84.62%) | 0.37 |
|  | Pelvic or abdominal pain | 3 (23.00%) | 6 (46.15%) | 0.25 |
|  | Nausea or vomiting | 1 (7.69%) | 2 (15.38%) | 0.99 |
| Post OPU | Pelvic or abdominal pain | 8 (61.54%) | 12 (92.31%) | 0.13 |
|  | Pain medication needed* | 10 (76.92%) | 12 (92.31%) | 0.62 |
|  | Nausea or vomiting | 2 (15.38%) | 2 (15.38%) | 0.48 |
|  | Bleeding | 7 (53.85%) | 9 (69.23%) | 0.62 |
|  | Fever | 0 (0.00%) | 0 (0.00%) | 1.00 |
|  | Regular menstrual cycle after a month | 11 (84.42%) | 10 (76.92%) | 0.99 |

\*Ibuprofen, paracetamol, and Statistical analysis performed via McNemar's Test

Since each participant underwent both stimulation protocols, we assessed how any given subjects’ symptoms differed between the two cycles. This allowed us to determine if a side effect was uniquely experienced during only a certain stimulation style or was commonly experienced or not experienced in both cycles. Importantly, analysis showed most side effects were only uniquely experienced during conventional stimulation treatment and not minimal stimulation such as breast swelling (2 cases), pelvic or abdominal pain (3 cases), nausea or vomiting (1 case). After OPU, a similar trend was found with specific participants only uniquely experiencing certain side effects after the conventional treatment and not after minimal stimulation such as abdominal or pelvic pain (4 cases), use of pain medications (2 cases), bleeding (3 cases), and non-recovery of menstrual cycle (1 case). Rarely was a symptom experienced only in the minimal stimulation and not in conventional stimulation, with only nausea or vomiting after OPU (2 cases), and bleeding (1 case) noted.

### Overall levels of pain in minimal and conventional stimulation treatments

Participants were asked about their overall level of pain post OPU. Similar to the findings observed in Data Set 1, a stark difference was observed in the “Severe” pain group with 38.46% of participants reported severe overall pain following conventional COS OPU, in contrast to the 7.69% of participants who indicated severe pain after undergoing minimal COS OPU (p=0.1336, McNemar’s Test, two-tailed) (Figure 4A). Importantly, for four subjects, minimal stimulation reduced their ‘severe’ pain experience in conventional stimulation to moderate or less. Overall, women experienced statistically significantly less pain after minimal COS treatment compared to conventional COS (p= 0.0078, Wilcoxon matched-pairs sign rank test, two-tailed) (Figure 4B). Importantly only one of thirteen women experienced more pain after the minimal stimulation versus conventional stimulation (4 ‘moderate’ versus 3 ‘mild’). In contrast, eight of thirteen participants experienced more pain after conventional versus minimal stimulation. This increase in pain levels is further mirrored by a trend towards increased requirement for pain medication in the conventional COS group (p=0.4795, McNemar’s Test, two-tailed) (Figure 4C).

**Figure 4.**
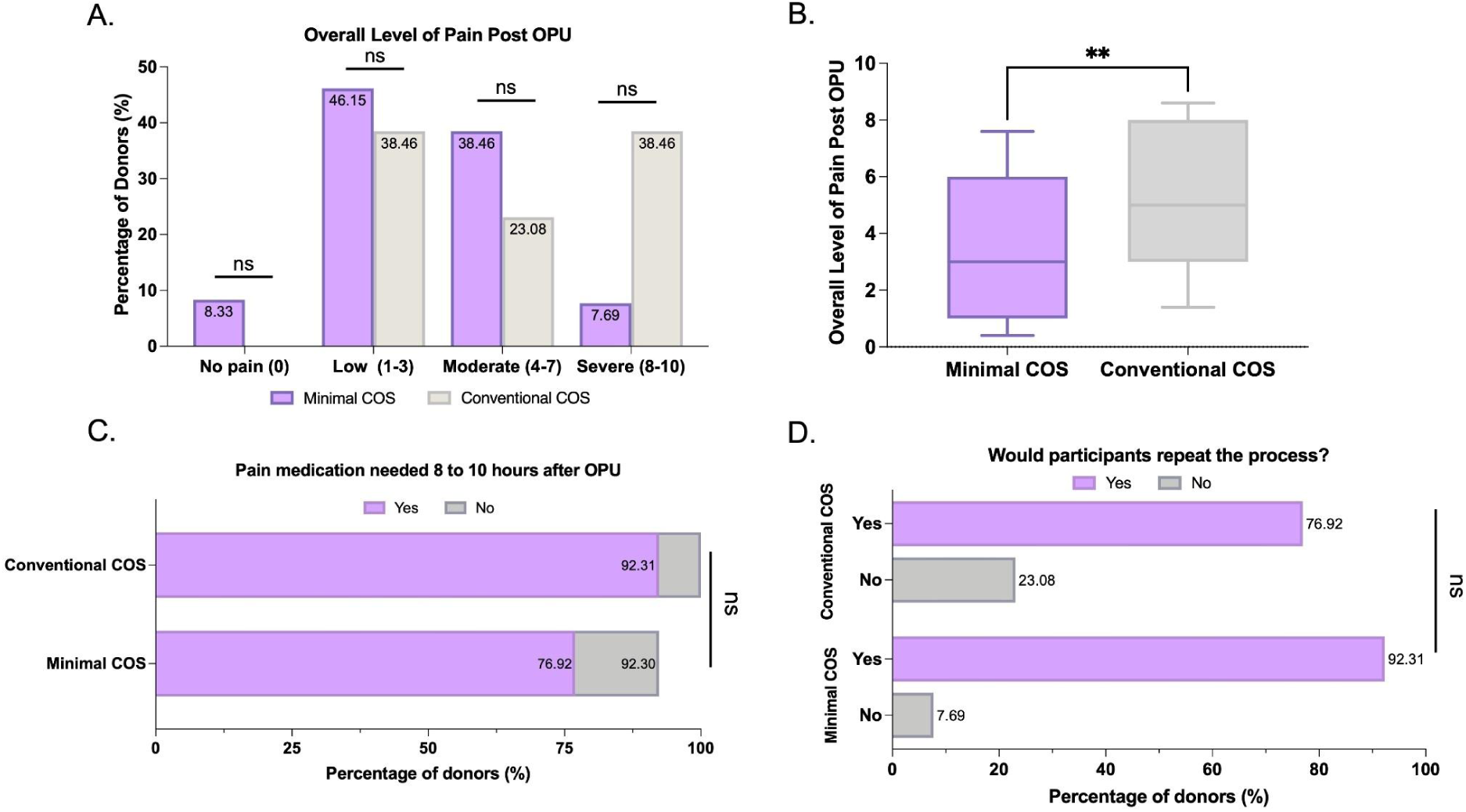
Pain experienced during minimal versus conventional ovarian stimulation and participant propensity for stimulation cycle repetition. Comparison of overall level of pain post oocyte-pick up (OPU) in the minimal controlled ovarian stimulation (COS) and conventional groups, expressed as a percentage for each pain level grouping. Statistical analysis was performed via McNemar’s test, two-tailed. ns, p>0.05 (B) Distribution of pain values in both conditions, scored on a scale of 0 to 10. Data is graphed as a box and whisker plot with median and 90% confidence interval. Statistical analysis was performed via Wilcoxon matched-pairs sign rank test, two-tailed,**, p=0.0078. (C) Percentage of subjects who needed pain medication 8 to 10 hours after OPU. Statistical analysis was performed via McNemar’s test, two-tailed. ns, p>0.05 (D) Percentage of subjects who would/would not recommend or repeat the minimal stimulation treatment. Statistical analysis was performed via McNemar’s test, two-tailed. ns, p>0.05.

### Propensity for cycle repetition minimal COS vs conventional COS

After undergoing both cycle types, participants in Data Set 2 were asked if they would repeat the stimulation and retrieval process for both minimal and conventional cycles. 92.31% of participants would recommend or repeat the minimal stimulation process again compared to a conventional cycle at 76.92% (Figure 4D).

### Comparison of efficacy and cost between minimal COS versus conventional COS treatment

A primary measure of efficacy in fertility treatment is the number of transferable embryos produced per cycle, which is directly correlated with treatment success odds (27). For this reason, we sought to provide an efficacy analysis of subjects that underwent minimal COS followed by IVM treatment, compared to conventional COS, to provide better context to overall treatment outcomes. We have recently published a comparative study evaluating the efficacy of our novel OSC-IVM treatment in maturing oocytes and generating healthy blastocysts embryos, compared to a traditional IVM treatment using MediCult IVM Media, in patients that underwent minimal COS (22). In that study the reported day 5 or 6 euploid blastocyst formation rate was 25% per COC in the OSC-IVM group and 11% per COC in the traditional IVM group, which we use here for our comparison of efficacy.

As that dataset only assessed the embryo formation outcomes of minimal COS plus IVM treatment, we sought a similar dataset for conventional IVF treatment as a further comparator beyond IVM. Recently, a non-inferiority US RCT was performed in predicted hyper responders for HP-hMG versus rFSH treatment (MEGASET-HR) (28). This study reported the subject attributes as well as oocyte retrieval and transferable blastocyst outcomes, which could be used to build a comparison against minimal COS plus IVM treatment. In that study, the reported excellent quality transferrable blastocyst formation rate was 19.9% per COC for HP-hMG and 17.6% per COC for rFSH treatment, which we use here for our comparison of efficacy.

A comparison between subject demographics of the two studies is shown in Table 4. As can be seen, the conventional IVF study arms had a significantly higher AMH and AFC, a similar BMI, a higher age, and significantly more gonadotropin stimulation and treatment length compared to the minimal COS plus IVM treatment. In order to provide a more equal comparison, we utilized the oocyte retrieval efficiency of the reported AFC for each study to simulate the number of COCs each treatment style would theoretically yield, and the reported transferrable blastocyst formation efficiency per COC to estimate the number of embryos that would be theoretically obtained in each treatment.

**Table 4:**
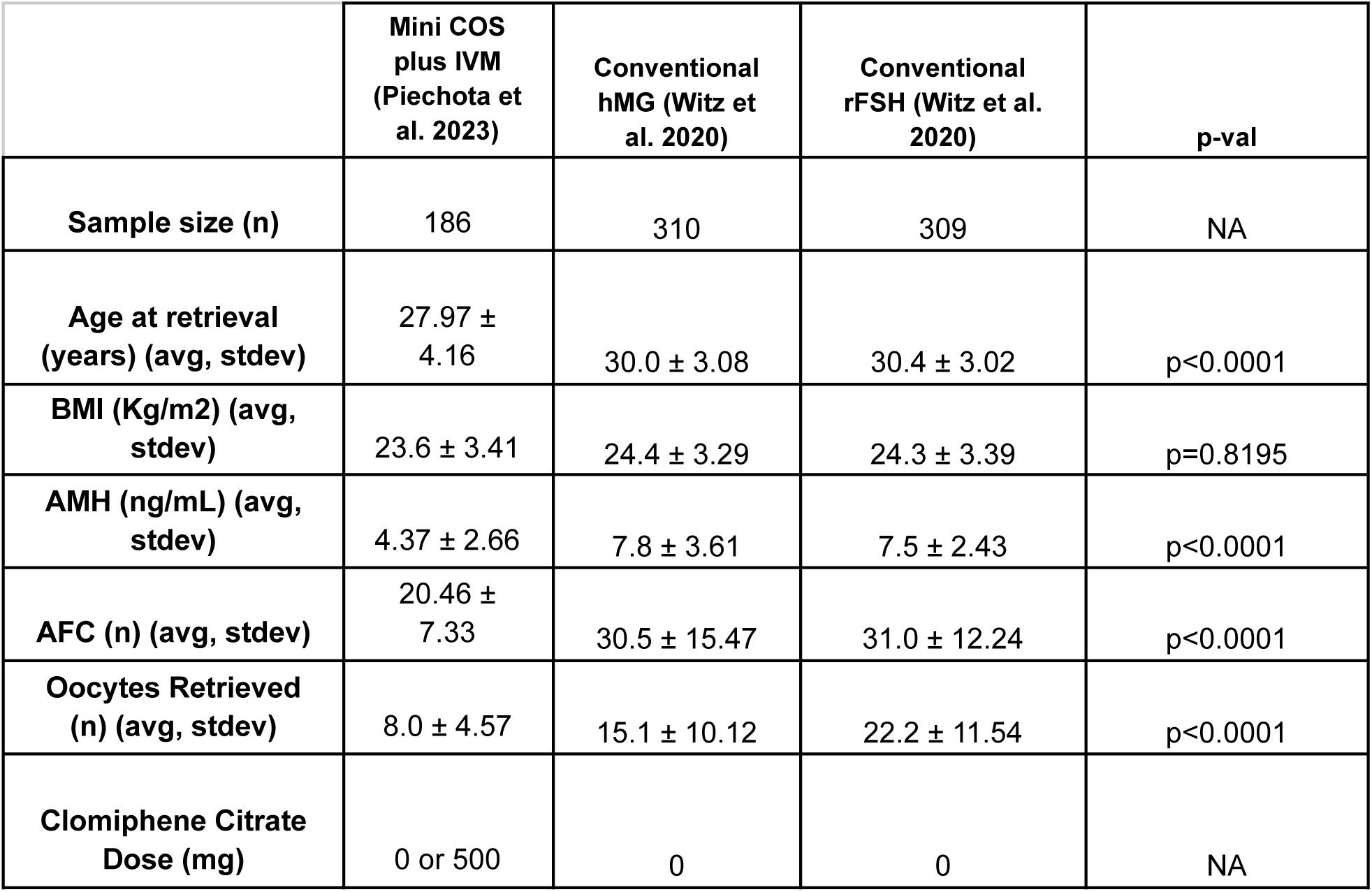
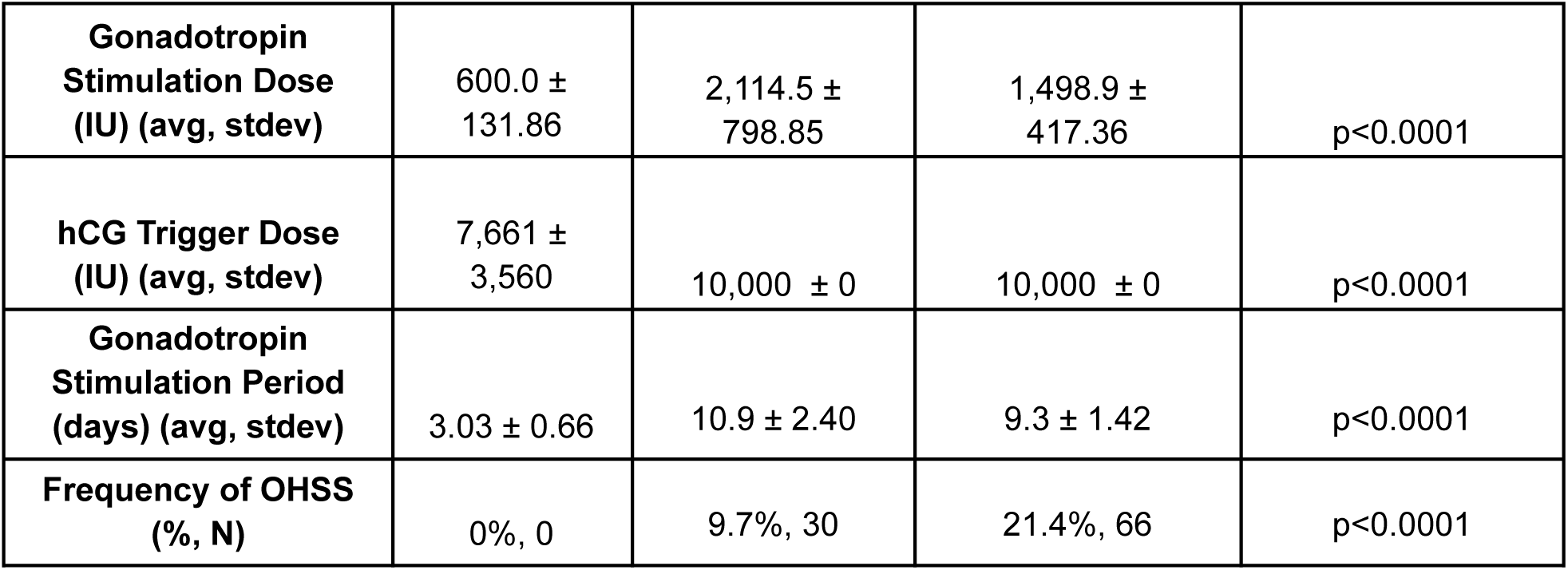
Comparison of demographic characteristics between comparator studies.

|  | Mini COS<br>plus IVM<br>(Piechota et<br>al. 2023) | Conventional<br>hMG (Witz et<br>al. 2020) | Conventional<br>rFSH (Witz et al.<br>2020) | p-val |
| --- | --- | --- | --- | --- |
| **Sample size (n)** | 186 | 310 | 309 | NA |
| **Age at retrieval**<br>**(years) (avg, stdev)** | 27.97 ±<br>4.16 | 30.0 ± 3.08 | 30.4 ± 3.02 | p<0.0001 |
| **BMI (Kg/m2) (avg,**<br>**stdev)** | 23.6 ± 3.41 | 24.4 ± 3.29 | 24.3 ± 3.39 | p=0.8195 |
| **AMH (ng/mL) (avg,**<br>**stdev)** | 4.37 ± 2.66 | 7.8 ± 3.61 | 7.5 ± 2.43 | p<0.0001 |
| **AFC (n) (avg, stdev)** | 20.46 ±<br>7.33 | 30.5 ± 15.47 | 31.0 ± 12.24 | p<0.0001 |
| **Oocytes Retrieved**<br>**(n) (avg, stdev)** | 8.0 ± 4.57 | 15.1 ± 10.12 | 22.2 ± 11.54 | p<0.0001 |
| **Clomiphene Citrate**<br>**Dose (mg)** | 0 or 500 | 0 | 0 | NA |
| **Gonadotropin Stimulation Dose (IU) (avg, stdev)** | 600.0 ± 131.86 | 2,114.5 ± 798.85 | 1,498.9 ± 417.36 | p<0.0001 |
| **hCG Trigger Dose (IU) (avg, stdev)** | 7,661 ± 3,560 | 10,000 ± 0 | 10,000 ± 0 | p<0.0001 |
| **Gonadotropin Stimulation Period (days) (avg, stdev)** | 3.03 ± 0.66 | 10.9 ± 2.40 | 9.3 ± 1.42 | p<0.0001 |
| **Frequency of OHSS (% , N)** | 0%, 0 | 9.7%, 30 | 21.4%, 66 | p<0.0001 |

For a hypothetical patient with an AFC of 25 (which was the midpoint between the reported AFC of the two studies), the projected embryo yield can be seen in Table 5. As can be seen, minimal COC plus OSC-IVM treatment theoretically yields a similar number of blastocysts to conventional IVF treatment using HP-hMG and rFSH (2.45 versus 2.47 and 3.15 respectively) while minimal COS plus traditional IVM treatment yields far less (1.08).

**Table 5:** Hypothetical efficacy analysis of IVM and IVF treatments.

|  | **Mini COS plus traditional IVM** | **Mini COS plus OSC-IVM** | **Conventional COS HP-hMG** | **Conventional COS rFSH** |
| --- | --- | --- | --- | --- |
| **Hypothetical Patient AFC** | 25 | 25 | 25 | 25 |
| **Hypothetical Oocyte Yield** | 9.8 | 9.8 | 12.4 | 17.9 |
| **Hypothetical Transferrable Blastocyst Yield** | 1.08 | 2.45 | 2.47 | 3.15 |

## Discussion

This study demonstrates that with minimal COS and IVM, individuals undergoing fertility treatment can avoid the risk of OHSS and reduce numerous common side effects associated with hormone treatment, such as breast swelling, nausea, fever, and pelvic or abdominal pain, compared to conventional stimulation cycles. In addition, by using minimal COS, the individuals recovered their regular menstrual cycles, showed high satisfaction levels, and the desire to repeat or recommend the stimulation process. In this study we found that pain post-OPU was reduced in the minimal stimulation despite the retrievals generally requiring more punctures due to needle flushing and the targeting of all visualized follicles. Additionally, in the minimal stimulation cohort both 17G and 19G needles were utilized, with no difference in pain level being attributed to needle diameter. These observations allowed us to conclude that the stimulation dosage and prolonged duration were the main contributors to pain experienced.

Factors such as youth, a history of OHSS, PCOS, and an elevated antral follicle count can contribute to OHSS and pronounced side effects associated with stimulation (33). In the case of PCOS patients, the high response of polycystic ovaries to gonadotropins creates a challenge in managing the response to COS, potentially resulting in OHSS and ovarian torsion in severe cases (34). For women with hormone-sensitive cancers, such as estrogen receptor-positive breast cancer, the prolonged ovarian simulations are also not recommended (35), as the administration of gonadotropins over 10–14 days to achieve multi-follicular development results in increased serum estradiol levels (36). In this retrospective study of minimal COS, no instance of ovarian torsion, OHSS, or hospitalization as a result of treatment was noted and acquisition of mature oocytes via IVM was accomplished with 2 to 4 days of minimal COS. Based on these findings, minimal COS and IVM may be an effective treatment strategy for improving patient safety during treatment for women at risk of OHSS and for women requiring abbreviated, low dose stimulation due to medical contraindication. It should be noted that a reduction in symptoms would also likely be expected in other forms of abbreviated hormonal stimulation and IVM when compared to conventional, and interestingly no significant difference in symptoms was noted in minimal stimulation cycles that utilized hCG triggers versus those that did not.

A significant challenge in assisted reproduction is establishing successful protocols for IVM for women who cannot or do not wish to undergo conventional COS, as the historically lower efficacy of IVM and lower relative oocyte retrieval rate are substantial hurdles to adoption (16–22,37). In previous studies, our group has demonstrated that using minimal stimulation protocols based on 2-4 days of FSH injections, or the usage of clomiphene citrate stimulation combined with an ovarian support cell-IVM treatment (OSC-IVM), allows obtaining healthy and mature eggs that can be cryopreserved or fertilized to generate to euploid day 5 or 6 blastocysts (22). These results show a potential application of minimal COS combined with IVM for high ovarian reserve patients for applications in oocyte cryopreservation and IVF. Other studies have likewise shown minimal COS plus IVM is an effective treatment option for PCOS and high ovarian reserve patients, resulting in quality embryo formation and healthy live births (20). Importantly, a recent study demonstrated that in patients where a large number of oocytes can be obtained, such as severe PCOS with AMH > 10 ng/ml, IVM treatment is non-inferior to IVF (38). In that study, one limitation of the IVM approach utilized (MediCult-IVM) was the overall low maturation rate (48.5%), which was overcome by application in patients with a sufficiently high number of COCs retrieved to yield enough embryos for transfer. It therefore stands to reason that improvement in the maturation and embryo formation rate could expand the applicability of IVM to beyond these severe PCOS cases while continuing to deliver improvement in treatment tolerability and risk of OHSS. A recent review likewise demonstrated minimal stimulation IVF, while yielding less oocytes compared to conventional COS IVF, obtains a similar number of high quality embryos and non-inferior birth outcomes while substantially reducing OHSS, stimulation dosage, and cost of care (39).

While the study data utilized as a retrospective dataset here was performed for basic research and not reproductive outcomes, the demonstration of improved maturation and embryo formation rates over the traditional IVM approach suggests this treatment could be used to broaden IVM application beyond its current common use cases. As well, the use of IVM in egg donors, social egg freezers, or when the cause of infertility is male infertility, in which the patient themselves are not necessarily infertile, presents an intriguing use case as these patients are not themselves being treated for infertility and therefore treatment strategies that reduce their risk or burden are important considerations when being subjected to medical intervention for IVF practice.

Minimal COS combined with IVM and IVF compared to conventional COS and IVF treatment is considered a less invasive, faster, and potentially less expensive treatment (40,41), making reproductive treatments more accessible for women. A recent study assessed costs in assisted reproduction, concluding that minimal COS was more cost-effective than conventional procedures due to significant savings from reduced hormone doses (14). Additionally, a reduction to 0 – 4 days of stimulation from 9–14 days in conventional COS means that women undergo significantly less monitoring in the form of ultrasound and bloodwork, often needing only one appointment with minimal COS and IVM compared to 3-6 appointments in conventional COS. Reduced monitoring needs in fertility treatment offer significant advantages for women, especially those facing travel challenges, leading to substantial time and cost savings for both patients and clinics, potentially enhancing treatment accessibility (23). When the embryo yield per cycle is hypothetically similar, such as the case for minimal COS plus OSC-IVM compared to conventional IVF, the cost per embryo using IVM is likely substantially less than conventional IVF.

While many studies have shown that historically minimal COS plus IVM resulted in inferior treatment success compared to conventional COS, recent advances in IVM and patient care have continued to improve these treatments, with embryo formation efficacy similar to conventional COS treatments and live birth rates only marginally below conventional IVF (19,22). It is widely known that success rates in IVF are highly variable depending on clinic, patient age, and infertility diagnosis and that women must often undergo multiple IVF rounds to achieve success, with the 2020 Society for Assisted Reproductive Technique (SART) analysis showing a 34% per cycle success rate in the United States (42). Patient drop-off or unwillingness to return after initial cycle failure represents a significant limitation in modern IVF, and the major physical, emotional, and financial burdens women undergo for treatment are important contributors to this drop-off. While less invasive options such as intrauterine insemination (IUI) may be successful for some, the success rates are extraordinarily low, with some studies showing a clinical pregnancy rate of 10% or less per cycle and pose inherent challenges in same-sex couples who wish to utilize reciprocal IVF treatment (43). This study shows that minimal COS plus OSC-IVM is highly tolerated and potentially similarly efficacious compared to conventional COS, with higher satisfaction and willingness to repeat. Furthermore, with overall cost and time savings, often requiring as little as three days, minimal COS plus IVM may present an ideal treatment choice as a first-line treatment for infertility or oocyte cryopreservation, particularly in patients with high ovarian reserve or a contraindication to conventional COS.

While the number of oocytes retrieved after minimal COS is often lower than conventional COS, wider adoption of the practice and improvements in retrieval techniques are likely to improve the rates. Likewise, the findings of this study suggest that given the tolerability of minimal COS plus IVM, patients may prefer to undergo two minimal COS plus IVM treatments versus one conventional COS treatment. Especially for women with PCOS, women undergoing fertility preservation before chemotherapy, and women resistant to gonadotropins, minimal COS and IVM represents an effective treatment option. Nevertheless, it’s important to remark that although IVM is no longer considered an experimental treatment by the European Society for Human Reproductive and Embryology (ESHRE) and American Society for Reproductive Medicine (ASRM), and over 5,000-6,000 live births have been documented worldwide to date (44,45), this technique still requires expertise gained through specific training and should be accompanied by appropriate counseling about expected results and informed consent (23).

### Study limitations

The minimal stimulation cycles analyzed in this study were conducted in fertility clinics as part of an IVM basic research study. This analysis is retrospective, which imposes limitations on the overall sample size and ability to properly power this study. For subjects in Data Set 1, the minimal COS cycles were not pairwise with the conventional cycles and represent nonoverlapping patients. The pain levels suffered in the conventional COS cycles were collected from oocyte subjects who voluntarily responded to the questionnaire at the clinic. In the case of Data Set 2, a highly valuable but limited sample size was available for women who underwent both minimal and COS stimulation protocols. A higher number of participants would be required to corroborate the differences observed between treatments.

Although the survey employed in this study was carefully designed, pain is a subjective experience, and individuals may perceive and describe it differently. Measurement scales were introduced to facilitate the assessment of pain and satisfaction levels. An overall limited number of questions were employed for analysis, and future studies could expand the analysis to a broader range of physical, emotional, and financial criteria. The inherently qualitative responses gathered here as well as reliance on self-reporting of symptoms could potentially result in recall bias, and a larger sample size and more quantitative metrics of outcomes are warranted to corroborate findings.

The efficacy analysis comparison to conventional IVF is limited by data availability. Caution should be drawn from the fact that minimal stimulation plus IVM and conventional IVF treatment outcomes compared here come from different studies and use a hypothetical approach, while may influence outcomes. Further, for the minimal COS plus IVM approaches discussed here, embryos were not generated for reproductive purposes, so the clinical pregnancy and live birth rates remain unknown, and further studies are warranted to compare the efficacy per cycle of these approaches.

## Conclusion

Subjects undergoing minimal hormonal stimulation protocol for IVM experienced few side effects, lower levels of pain after retrieval, and high satisfaction levels with the procedure. Therefore, reducing gonadotropin levels with shorter stimulation protocols followed by successful IVM techniques could be preferential for many women as it means lower cost and side effects, as well as a possible treatment for women for whom prolonged conventional stimulation is contraindicated.

## Data Availability

Data Sharing Statement: All data needed to evaluate the conclusions in the paper are present in the paper and supplementary tables and figures.

## Acknowledgements

The authors thank the dedicated support and work of the embryology and staff teams at Spring Fertility New York, Extend Fertility, and Ruber Juan Bravo University Hospital, Eugin Group for coordinating and managing the collaborative study.

## Contributors

M.M., S.P., M.F., A.G. analyzed all subject data. T.S., A.S., and E.G. coordinated and collected subject response data. S.K. performed statistical analysis of data. F.B., and B.P. edited manuscript and assisted in data analysis. J.K., B.A., V.C., D.O., and P.K. performed all donor retrievals and monitoring. C.C.K. designed, supervised, and coordinated the study. M.M., S.P., and C.C.K wrote the manuscript with significant contribution from all authors.

